# Incident Atrial Fibrillation and Flutter in Patients with Pulmonary Arterial Hypertension: Influence of Right Ventricular Dilatation and Reduced Right Atrial Function

**DOI:** 10.1101/2025.02.05.25321674

**Authors:** Danny Mohama, Pitchaya Worapongsatitaya, Bettia Celestin, Felipe Kazmirczak, Shadi P. Bagherzadeh, Kurt W. Prins, Sasha Z. Prisco, E. Kenneth Weir, Stephen L. Archer, Roham Zamanian, Francois Haddad, Thenappan Thenappan

## Abstract

**Background:** The relationship between right atrial (RA) structural and functional remodeling and risk of atrial fibrillation/flutter (AF/AFL) in pulmonary arterial hypertension (PAH) is understudied. This is important due to the prognostic implications of AF/AFL in PAH.

**Methods:** In a multicenter cohort study comprised of 326 PAH patients with no prior history of AF/AFL, we evaluated how RA structure and function, as determined by echocardiography, were associated with AF/AFL. We calculated the incident rate (IR) of AF/AFL using time to event analysis. Cox proportional hazards analyses defined factors associated with incident AF/AFL and Harrell’s C-statistics compared the ability of different variables to predict incident AF/AFL. Survival decision tree and restricted cubic spline analyses identified thresholds associated with incident AF/AFL.

**Results:** In the combined PAH cohort, the mean age was 51±15 years, 79% were female, and the mean REVEAL lite score was 7.4±2.9. Over a median follow-up of 6.1 years, 56 patients (17.1%) developed AF/AFL with an IR of 25.3 cases (95% CI: 19.5 - 32.8) per 1000 person-year. On multivariable Cox proportional hazards analysis, every 5% decrease in RA emptying fraction (RAEF, HR: 1.38, 95% CI: 1.03 – 1.86, *p*=0.030) and one centimeter increase in right ventricular (RV) basal diameter (HR: 1.55 95% CI: 1.18 – 2.05, *p*=0.002) were independently associated with 38% and 56% increased hazards of incident AF/AFL, respectively. The C-statistics of RAEF and RV basal diameter to predict incident AF/AFL were 0.62 and 0.65, respectively. Survival decision tree and restricted cubic spline analyses identified informative thresholds for RAEF at <17 and 45% and for RV basal diameter at ≥5.4 cm for increased hazards of incident AF/AFL.

**Conclusion:** Lower RAEF and higher RV basal diameter are associated with increased risk of incident AF/AFL in PAH patients. These data could help identify PAH patients at risk for AF/AFL.

## Introduction

Pulmonary arterial hypertension (PAH) is characterized by remodeling of the distal pulmonary arteries leading to an increase in pulmonary artery pressure and pulmonary vascular resistance(1). PAH is chronic, progressive, and eventually leads to premature death due to right heart failure(1). The right ventricle (RV) initially adapts to the increased afterload in the pulmonary circulation by hypertrophy and increased contractility, which maintains stroke volume(2). Eventually, the RV dilates, which maintains stroke volume by increasing the RV end-diastolic volume(2). These adaptive and maladaptive RV changes in turn lead to right atrial (RA) structural remodeling and mechanical dysfunction(3).

While left atrial (LA) structural and functional remodeling is clearly associated with an increased risk of atrial fibrillation and atrial flutter (AF/AFL) in patients with left heart disease(4), little is known about the incidence and the risk of AF/AFL due to RA structural and functional remodeling in PAH patients (5–8). This is relevant as the presence of AF/AFL is associated with increased mortality in PAH(5–8). A better understanding of the determinants of AF/AFL in PAH patients can potentially lead to better monitoring strategies, earlier diagnosis, and improved outcomes.

Therefore, in this study, we sought to determine the incidence and risk of AF/AFL in patients with PAH based on different RA structural and functional parameters assessed by two-dimensional echocardiography (2DE).

## Methods

### Study Population

We studied adult PAH patients with no prior history of AF/AFL in two PAH cohorts (Minnesota Pulmonary Hypertension Repository and Stanford Pulmonary Hypertension Registry). The Minnesota Pulmonary Hypertension Repository is a single center registry that enrolls patients treated for pulmonary hypertension at the University of Minnesota Pulmonary Hypertension Clinic since March 2014. Patients who were diagnosed before March 2014 were entered retrospectively. Data were collected by chart review and entered using an internet-based electronic data-capture system. Informed consent was obtained from every participant in the repository. Minnesota Pulmonary Hypertension Repository collected baseline incident data at the time of diagnosis. The University of Minnesota institutional review board approved the Minnesota Pulmonary Hypertension Repository (IRB#1310M44485). The Stanford cohort consists of adult PAH patients, both incident and prevalent, followed at Vera Moulton Wall Center for Pulmonary Vascular Disease at Stanford Hospital between January 2002 and 2021. The Stanford University Institutional Review Board approved this protocol conducted under the CardioShare protocol (IRB#25673).

PAH was defined as mean pulmonary arterial pressure (mPAP) > 20 mmHg, pulmonary capillary wedge pressure (PCWP) ≤ 15 mmHg, and pulmonary vascular resistance (PVR) > 2 Wood units at rest (9). Patients with other World Health Organization (WHO) categories of pulmonary hypertension (PH) were excluded by clinical evaluation and objective tests, including pulmonary function tests, ventilation-perfusion (V/Q) scan, and pulmonary angiography. Patients with obstructive lung disease, diagnosed by reduced expiratory flow rates (forced expiratory volume in one second [FEV1]/forced vital capacity [FVC] <75% predicted), more than mild interstitial lung disease, diagnosed by reduced total lung capacity <60%, and chronic thromboembolic pulmonary hypertension diagnosed by high probability V/Q scan or pulmonary angiography were excluded. In addition, we excluded patients who had a prior history of AF/AFL at the time of enrollment.

### Echocardiographic Assessment

Echocardiographic images were acquired using Hewlett Packard Sonos 5500 or Philips IE33 ultrasound systems with a 3.5-MHz multiphase-array probe according to the American Society of Echocardiography guidelines for chamber quantification(9). Echocardiographic analysis was performed off-line by two experienced readers with advanced training in cardiovascular imaging and blinded to clinical and hemodynamic data (FK and BC). Measurements of RA and RV dimensions were conducted at end-diastole and end-systole defined by largest and smallest chamber size. Comprehensive analysis of the cardiac chambers was performed including measurement of right atrial linear dimensions, right atrial area, right atrial volume, right atrial emptying fraction (RAEF), RV basal diameter, right ventricular fractional area change (RVFAC), severity of tricuspid regurgitation, left atrial volume, and left ventricular ejection fraction(9). RA volume was calculated based on the single plane area-length method(9). Area and volume measurements were indexed to body surface area. RA emptying fraction (RAEF) was defined as (RA maximum volume – RA minimum volume)/RA maximum volume. RV basal diameter was measured on RV focused apical 4-chamber view at end diastole. RVFAC was defined as (RV end diastolic area – RV end systolic area)/RV end diastolic area.

### Clinical Covariates

We analyzed the following baseline demographic and clinical characteristics including age, sex, body mass index (BMI), etiology of PAH, WHO functional class, 6-minute walk distance (6MWD), serum N-terminal fragment of pro-Brain Natriuretic Peptide (NT-proBNP) levels, Registry to Evaluate Early and Long-term outcomes in PAH (REVEAL) lite score, and hemodynamics from right heart catheterization (RHC). REVEAL lite score was calculated for all participants as described previously (11). Hemodynamic measurements were evaluated from RHC data including mean right atrial pressure (mRAP), systolic, diastolic, and mean pulmonary artery pressures, PCWP, cardiac output, cardiac index, and PVR.

### Outcomes

The primary outcome was incident AF/AFL. Incident AF/AFL was obtained by detailed review of the electronic health records for each participant. The date of onset of the incident AF/AFL was recorded.

### Statistical Analysis

Continuous variables were reported as mean ± standard deviation or median (interquartile range), and categorical variables were presented as frequency and percentage unless indicated. We used unpaired t-test or Wilcoxon-Mann Whitney test to compare continuous variables and χ2 test or Fisher Exact test for comparing categorical variables. We calculated the incident rate (IR) of AF/AFL using time to event analysis with entry into the study defined as the date of baseline echocardiographic assessment. The primary outcome was incident AF/AFL. Patients were censored if they were lost to follow up, had lung transplantation, died, or at study completion. We used univariable and multivariable Cox’s proportional hazards analyses to identify the determinants of incident AF/AFL. As there is high collinearity between different RA parameters, we created a multivariable model for each RA parameter adjusting for other univariable predictors of incident AF/AFL including RVFAC, RV basal diameter, severity of tricuspid regurgitation, mean RA pressure, and REVEAL lite risk categories. We did not include 6MWD in the multivariable models as it is already incorporated into the calculation of REVEAL lite score. All hazard ratios were scaled to standard deviation to provide better comparison. We used receiver operative characteristic (ROC) analysis and Harrell’s C-statistics to compare the ability of different parameters to predict incident AF/AFL. We determined the optimal cutoff values associated with increased hazards of incident AF/AFL by survival decision tree and non-linear Cox regression analyses. We compared the probability of incident AF/AFL between groups using Kaplan-Meier analysis, log rank test, and incident rate ratio (IRR).

Survival tree and non-linear Cox regression survival analyses were conducted with R (version 4.4.1). Survival tree analysis used the rpart package to assess time-to-event outcomes, with key predictors visualized using rpart.plot. Non-linear Cox regression analysis was performed with the survival and rms packages, employing restricted cubic splines for continuous predictors. Model performance was evaluated using the C-statistic, reported with 95% confidence intervals. Hazard ratios and their confidence intervals were visualized using ggplot2 for clarity on predictor effects. All other analyses were performed using STATA (version IC). A p-value <0.05 was considered statistically significant.

## Results

### Patient Characteristics

We identified 400 patients with PAH who had baseline echocardiographic and hemodynamic data in the Minnesota Pulmonary Hypertension Repository and Stanford University Pulmonary Hypertension Registry. Of the 400 PAH patients, 74 patients were excluded as they did not meet hemodynamic criteria for PAH or had prior AF/AFL (**Figure 1**). The remaining 326 patients formed our study cohort, of which 141 patients were from the Minnesota Pulmonary Hypertension Repository and 185 patients were from Stanford University’s Pulmonary Hypertension Registry (**Figure 1**).

**Figure 1.**
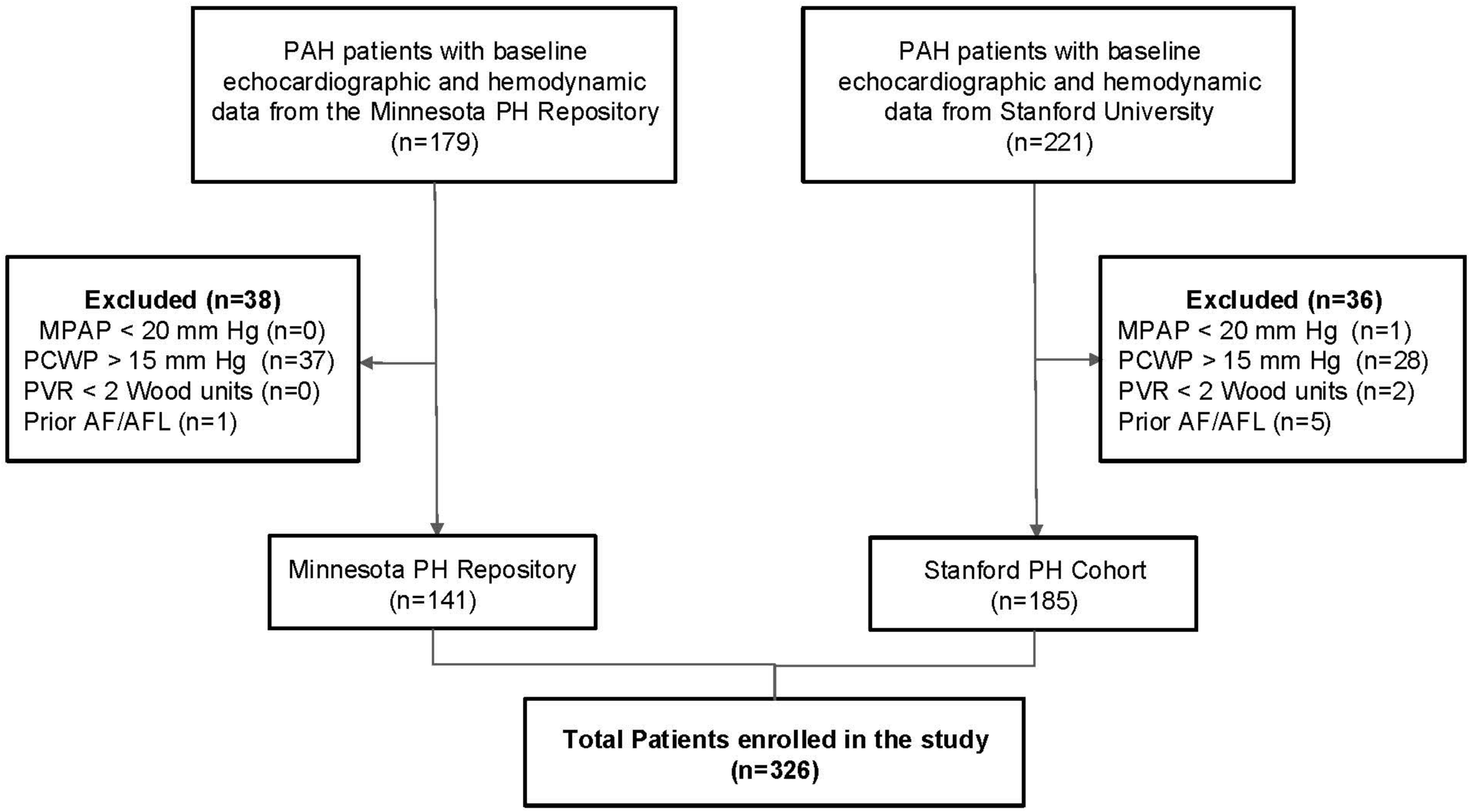
Strobe Diagram of the Study Cohort We identified 400 patients with PAH who had baseline echocardiographic and hemodynamic data in the Minnesota Pulmonary Hypertension Repository and Stanford University Pulmonary Hypertension Registry. Seventy-four patients were excluded as they did not meet hemodynamic criteria for PAH or had AF/AFL. The remaining 326 patients formed our study cohort, of which 141 patients were from the Minnesota Pulmonary Hypertension Repository and 185 patients were from Stanford University Pulmonary Hypertension Registry. PH – pulmonary hypertension, PAH – pulmonary arterial hypertension, mPAP – mean pulmonary artery pressure, PVR – pulmonary vascular resistance, PCWP – pulmonary capillary wedge pressure, and AF/AFL – atrial fibrillation and/or atrial flutter.

**Table 1** delineates the baseline clinical, hemodynamic, and echocardiographic characteristics of the study cohort. The mean age was 51±15 years, 258 (79%) were female, and 71.5% had WHO functional class III or IV symptoms. The most common etiology was connective tissue disease followed by idiopathic PAH, and drug-and toxin-related PAH. Patients, on average, had severe PAH with a mPAP of 48±13 mmHg, PVR of 11.5±6.3 WU, PCWP of 10±3 mm Hg, and cardiac index of 2.1±0.7 L/min/m^2^. The mean REVEAL lite score was 7.4±2.9 and mean 6MWD was 337±155 meters. The median serum NT-proBNP level was 789 (IQR: 231–2495) pg/ml.

**Table 1:**
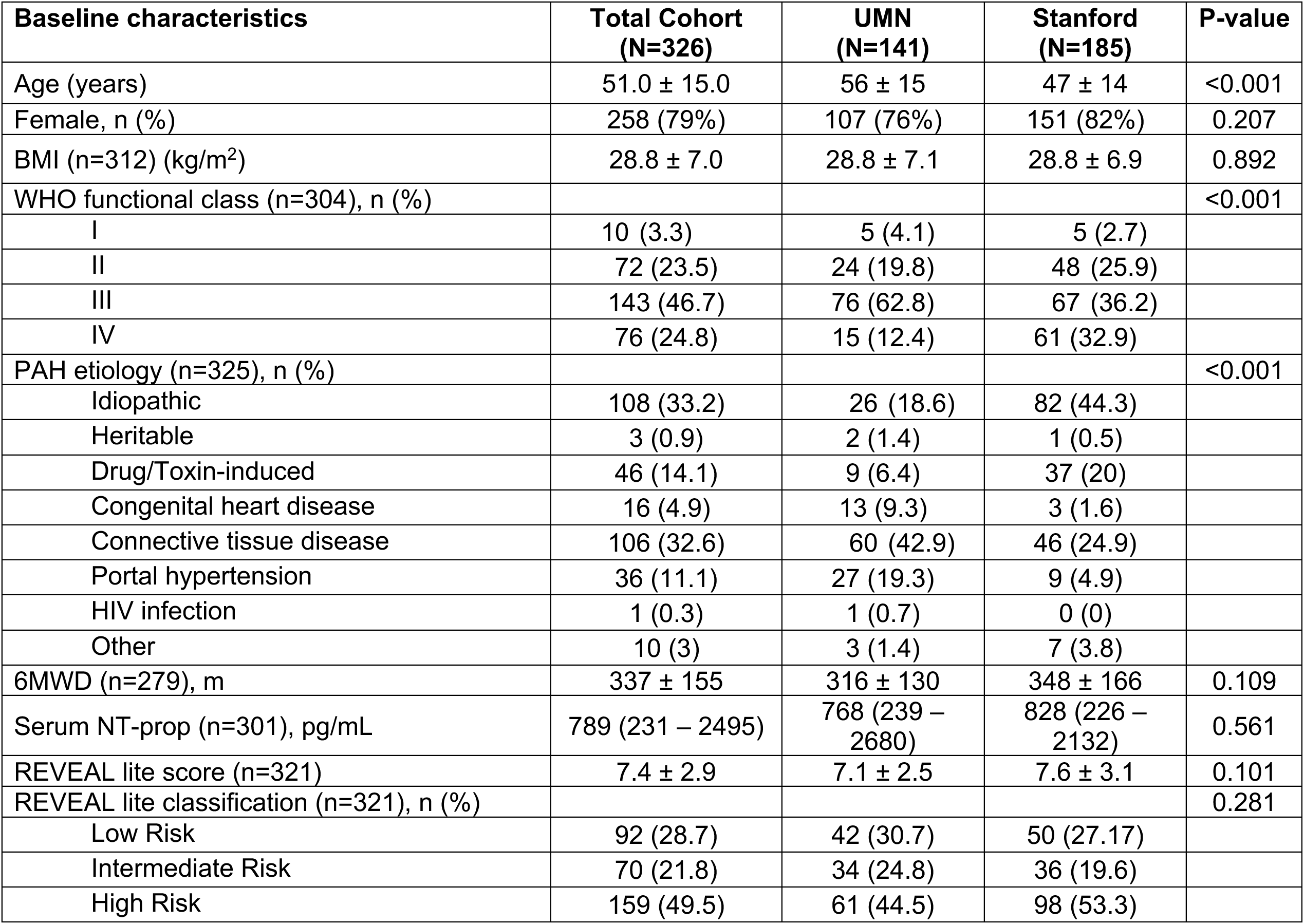
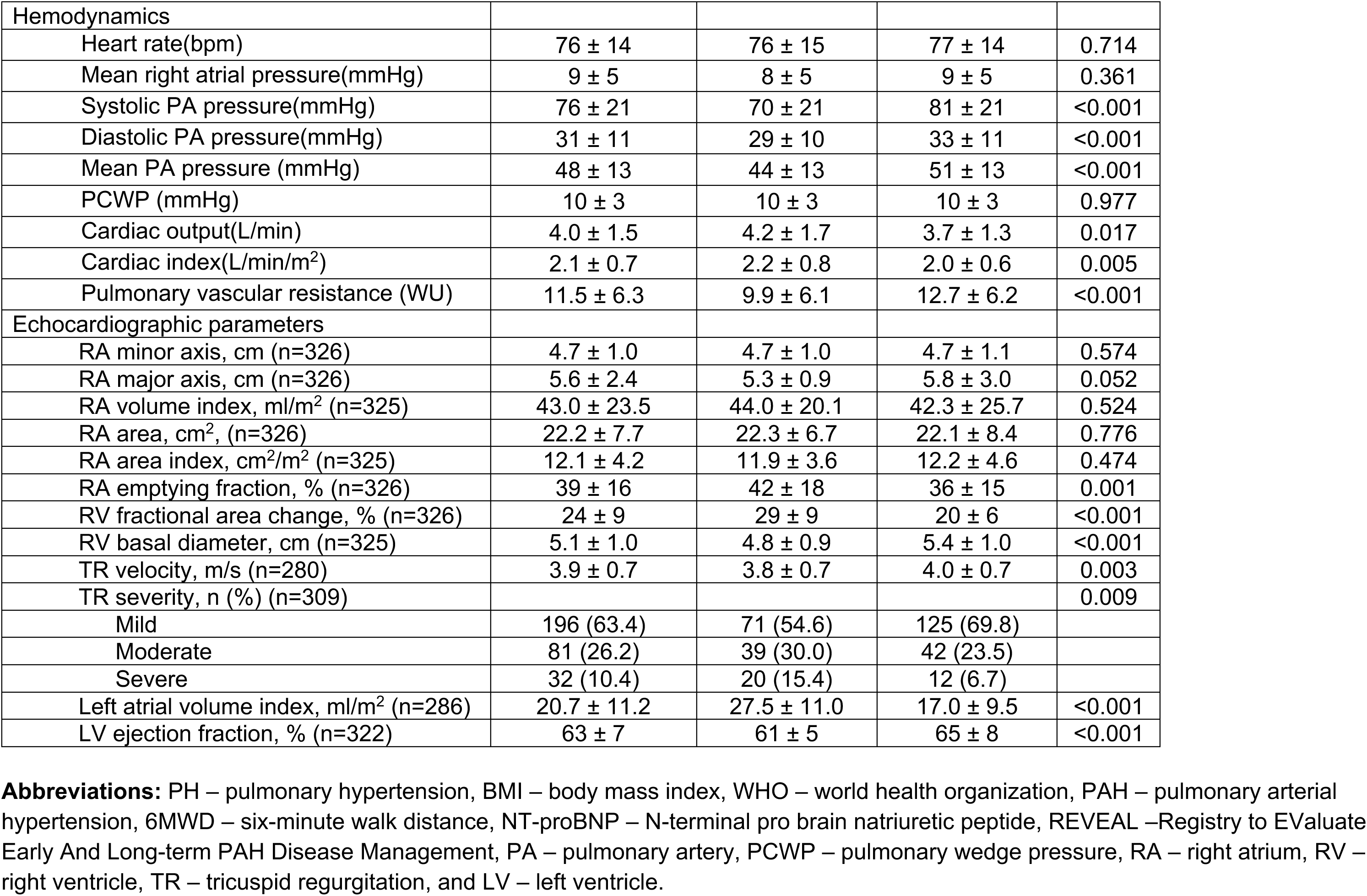
Baseline Demographic, Clinical, and Hemodynamic Characteristics of the Study Cohort.

The mean RA major axis diameter was 5.6±2.4 cm (normal < 5.3 cm), the RA minor axis diameter was 4.7±1.0 cm (normal <4.4 cm), RA area was 22.2±7.7 cm^2^ (normal < 18 cm^2^), RA area index was 12.1±4.2 cm^2^/m^2^ (normal < 8.5 cm²/m²), RA volume index was 43.0±23.5 ml/m^2^ (normal men: 25±7 and female: 21±6 ml/m^2^), and mean RAEF was 39±16% (normal 45-50%). The mean RVFAC was 24±9% and RV basal diameter was 5.1±1.0 cm. Nearly one third of the patients (36.6%) had moderate or severe tricuspid regurgitation. The mean left atrial volume index and left ventricular ejection fraction were 20.7±11.2 ml/m^2^ and 63.3 ± 7.0%, respectively.

Compared to the patients in the Minnesota Pulmonary Hypertension registry, patients in the Stanford Pulmonary Hypertension registry were younger, less often had WHO functional class III and IV symptoms, were more likely to have idiopathic PAH, had severe PAH with higher mPAP, higher PVR, and lower cardiac index, and had more adverse RA and RV remodeling with lower RAEF, lower RVFAC, higher RV basal diameter, less severe tricuspid regurgitation, lower left atrial volume index, and higher left ventricular ejection fraction (**Table 1**).

### Incidence, Prognostic Significance, and Determinants of Atrial Fibrillation and Atrial Flutter

Over a median follow-up of 6.1 years, fifty-six patients (17.1%) developed AF/AFL with an incidence rate of 25.3 cases (95% CI: 19.5 - 32.8) per 1000 person-year. Of the 17.1% who developed incident AF/AFL, 87.5% had AF and 12.5% AFL. Incident AF/AFL was associated with a 2.1-fold increased hazard of death or transplant (HR: 2.1, 95% CI: 1.3–3.4, *p* = 0.001, **Supplemental Figure 1**). Survival rates after the onset of incident AF/AFL were significantly lower, with 1-, 3-, and 5-year survival rates of 73%, 57%, and 48%, respectively, compared to 91%, 80%, and 75% in patients without incident AF/AFL (*p* = 0.001).On univariable analysis, incident AF/AFL was significantly associated with REVEAL risk score, REVEAL high risk status, 6MWD, severity of tricuspid regurgitation, RVFAC, RV basal diameter, mean RA pressure, RA major axis diameter, RA minor axis diameter, RA area and area index, RA volume index, and RAEF (**Table 2**).

**Table 2:**
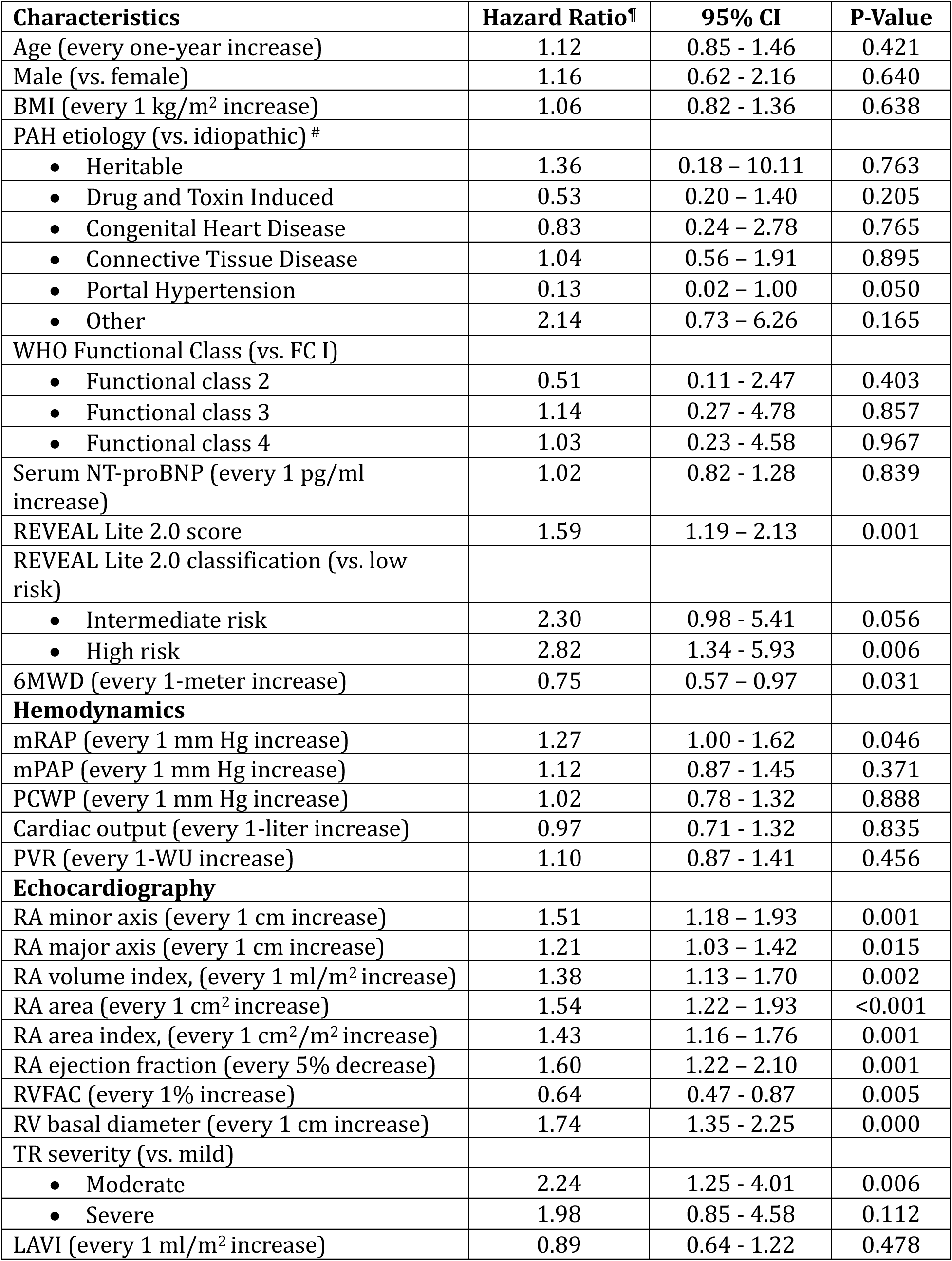
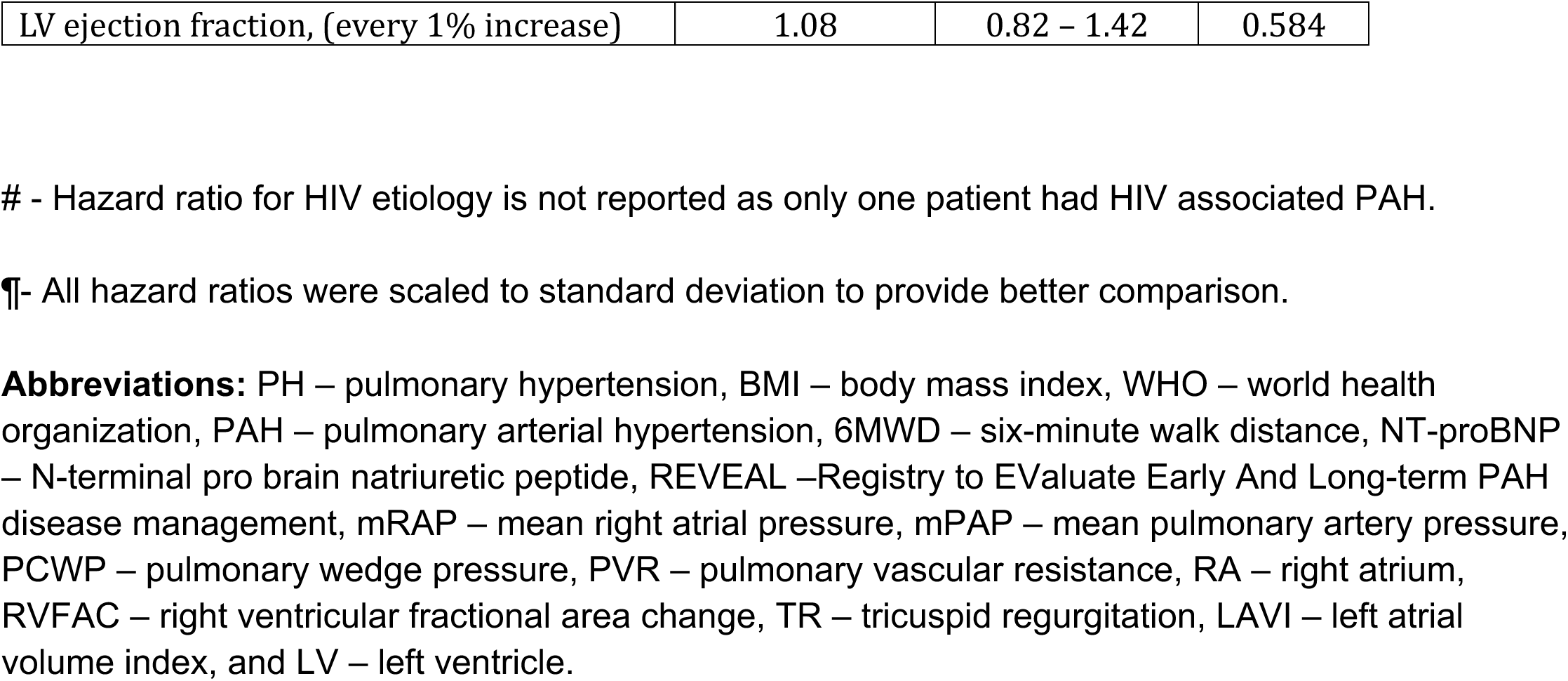
Univariable Predictors of Incident Atrial Fibrillation and Flutter in Pulmonary Arterial Hypertension.

To identify whether RA structural and functional remodeling is independently associated with incident AF/AFL, we created a multivariable model for each RA parameter adjusting for other univariable predictors of incident AF/AFL including RVFAC, RV basal diameter, severity of tricuspid regurgitation, mean RA pressure, and REVEAL risk status. On these multivariable models, RAEF and RV basal diameter were the only independent predictors of incident AF/AFL (**Table 3**). Every 5% decrease in RAEF (HR: 1.38, 95% CI: 1.03-1.86, *p*=0.030) and each one cm increase in RV basal diameter (HR: 1.55, 95% CI: 1.18-2.05, *p*=0.002) were independently associated with 38% and 55% increased hazards of incident AF/AFL, respectively. When we included REVEAL lite risk score in the model as opposed to REVEAL risk status, RV basal diameter (HR: 1.52, 95% CI: 1.16 – 2.01, *p*=0.003) and REVEAL risk score (HR: 1.50, 95%CI: 1.09-2.07, *p*=0.012) were the independent predictors of incident AF/AFL.

**Table 3:** Multivariable Predictors of Atrial fibrillation and flutter in PAH Categorized by Right Atrial Structural and Functional Parameters. Each Model is adjusted for REVEAL risk status, Right Ventricular fractional Area Change, Right Ventricular Basal Diameter, Severity of Tricuspid Regurgitation, and Mean Right Atrial Pressure.

| Characteristics | Hazard ratio<sup>¶</sup> | 95% CI | P-Value |
| --- | --- | --- | --- |
| **Model 3A: RA volume index** |  |  |  |
| • RV basal diameter<br>(for every 1 cm increase) | 1.68 | 1.29 - 2.17 | <0.001 |
| **Model 3B: RA area index** |  |  |  |
| • RV basal diameter<br>(for every 1 cm increase) | 1.68 | 1.29 - 2.17 | <0.001 |
| **Model 3C: RA area** |  |  |  |
| • RV basal diameter<br>(for every 1 cm increase) | 1.68 | 1.29 - 2.17 | <0.001 |
| **Model 3D: RAEF** |  |  |  |
| • RV basal diameter<br>(for every 1 cm increase) | 1.55 | 1.18 - 2.05 | 0.002 |
| • RAEF<br>(for every 5% decrease) | 1.38 | 1.03 – 1.86 | 0.030 |
| **Model 3E: RA minor diameter** |  |  |  |
| • Right ventricular basal diameter<br>(for every 1 cm increase) | 1.68 | 1.29 – 2.17 | <0.001 |
| **Model 3F: RA major diameter** |  |  |  |
| • RV basal diameter<br>(for every 1 cm increase) | 1.68 | 1.29 – 2.17 | <0.001 |

¶- All hazard ratios were scaled to standard deviation to provide better comparison.
**Abbreviations:** RA – right atrium, RV – right ventricle, RAEF – right atrial emptying fraction, and REVEAL - Registry to EValuate Early And Long-term PAH disease management.

### Comparison of the Ability to Predict Incident AF/AFL and Identification of Optimal Threshold

We then compared the ability of both RV basal diameter and RAEF to predict incident AF/AFL. There was no significant difference between RV basal diameter and RAEF in predictive capacity, with an area under the ROC curve of 0.67 and 0.65 (*p*=0.569), respectively (**Figure 2**). The Harrell’s C-statistics for predicting incident AF/AFL was 0.65 and 0.62 for RV basal diameter and RAEF, respectively.

**Figure 2.**
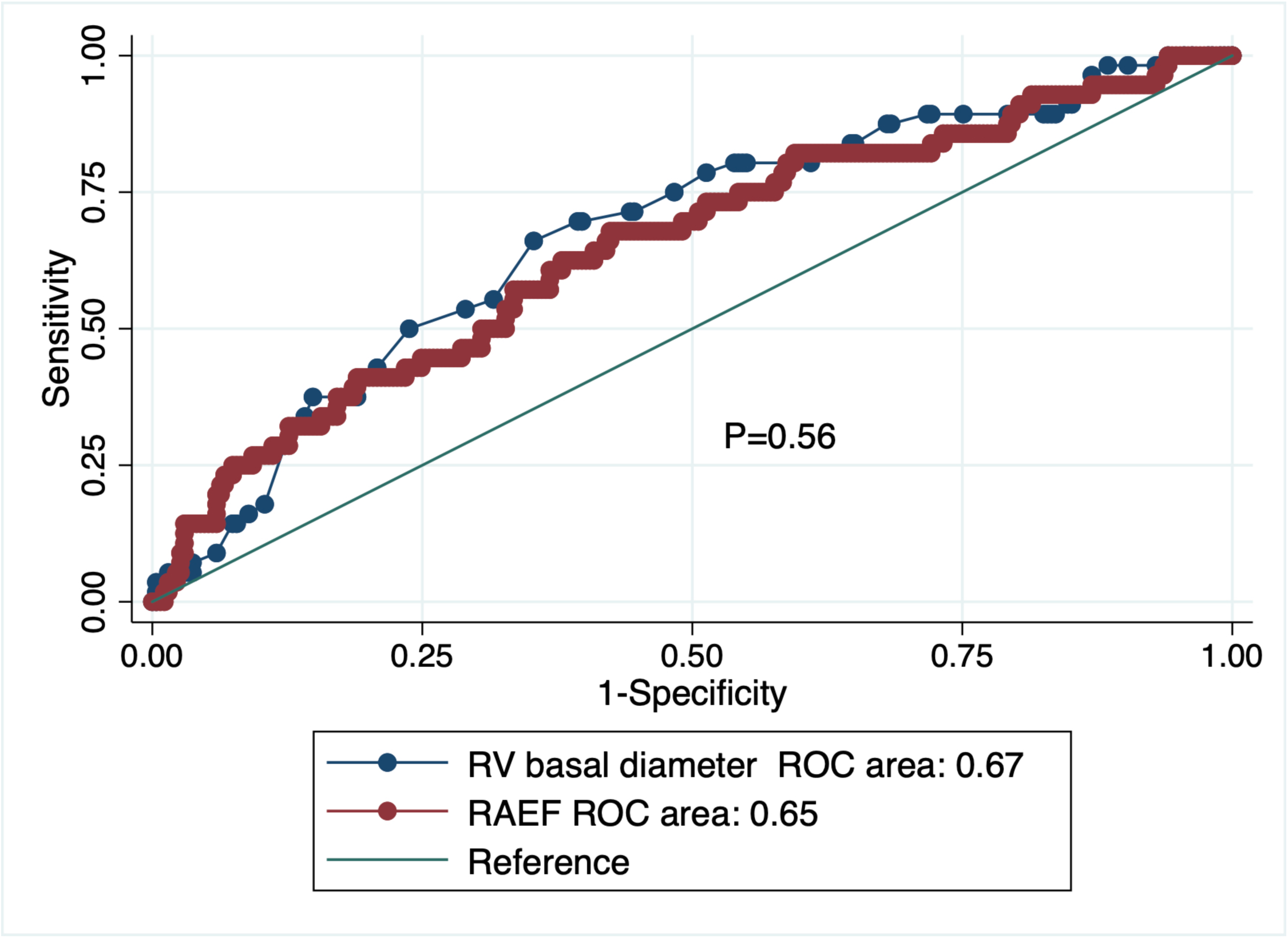
Receiver Operating Curve Comparing the Predictive Ability of Right Atrial Emptying Fraction and Right Ventricular Basal Diameter for Incident Atrial Fibrillation and Atrial Flutter There was no significant difference in the area under the curve for right atrial emptying fraction and right ventricular basal diameter to predict incident atrial fibrillation and atrial flutter.

On multivariable survival decision tree analysis including both RV basal diameter and RAEF, RV basal diameter ≥5.4 cm had a stronger ability for predicting incident AF/AFL compared to RAEF, with RV basal diameter at the root of the tree (**Figure 3A**). In patients with RV basal diameter ≥5.4 cm, even a mild reduction in RAEF (<45%) was associated with a 2.1-fold increased hazards of incident AF/AFL (**Figure 3A)**. In contrast, in patients with RV basal diameter <5.4 cm, only a significant reduction in RAEF (to values <17%) was associated with 1.8-fold increased hazards of incident AF/AFL (Figure 3A). Non-linear Cox regression analysis including both RV basal diameter and RAEF identified similar thresholds (**Figure 3B and 3C**).

**Figure 3.**
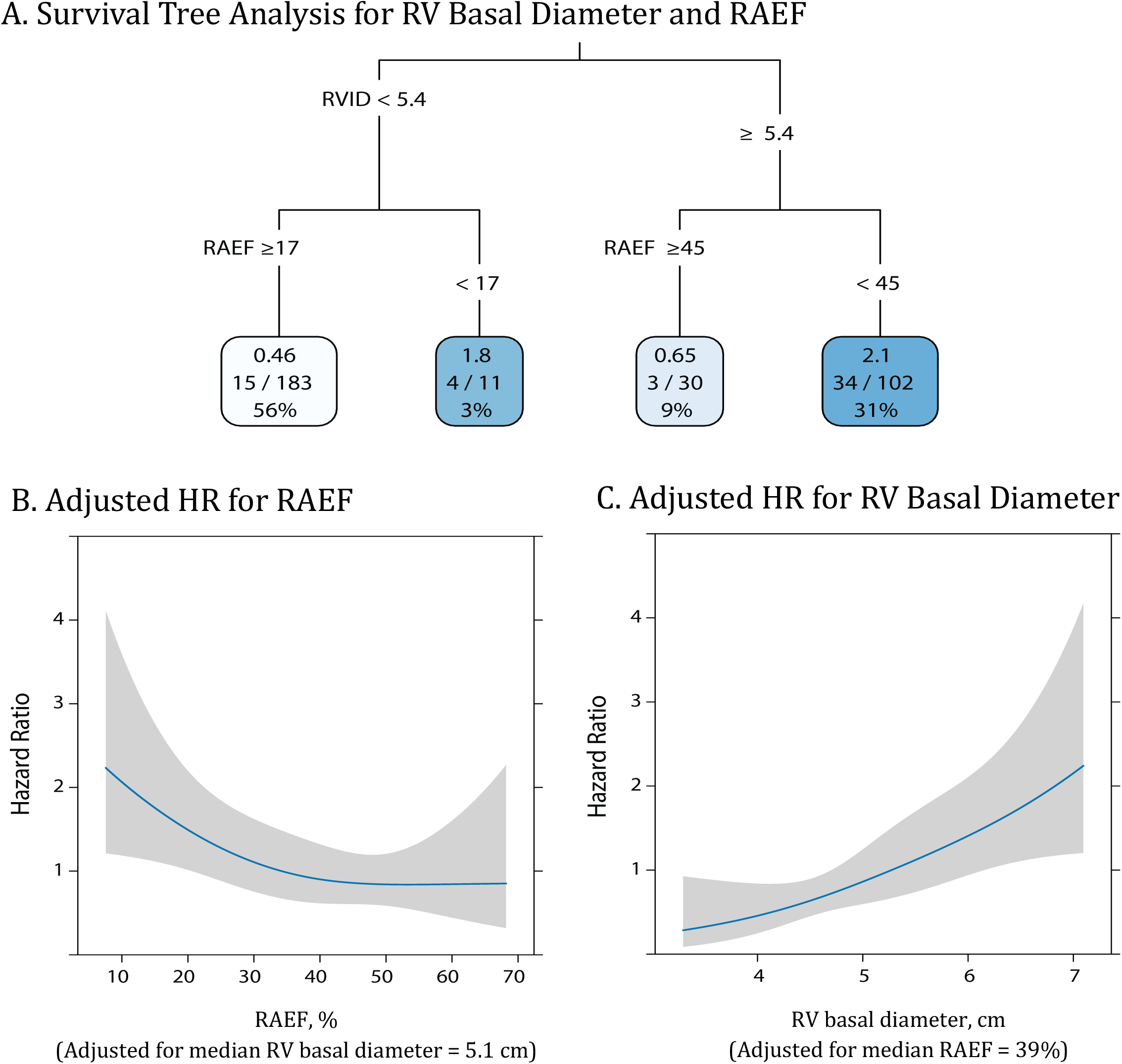
Multivariable Survival Decision Tree and Non-linear Restricted Cubic Spline Analyses for Identifying Thresholds of RV Basal Diameter and RAEF Associated with Incident Atrial Fibrillation and Flutter in PAH (A) Survival tree analysis identifies RV basal diameter ≥5.4 cm as the strongest predictor of incident AF/AFL. In patients with RV basal diameter ≥5.4 cm, even a mild reduction in RAEF (<45%) is associated with a 2.1-fold increased hazards of incident AF/AFL. In contrast, in patients with RV basal diameter <5.4 cm, only a significant reduction in RAEF (<17%) is associated with 1.8-fold increased hazards of incident AF/AFL (Figure 3A). (B) Nonlinear restricted cubic spline analyses adjusted for median RV basal diameter, shows that the risk of incident AF/AFL increases when the RAEF is below 45%, but the risk increases significantly when the RAEF is <17%. (C) Similarly, nonlinear restricted cubic spline analyses adjusted for median RAEF demonstrates that the risk of AF/AFL increases when the RV basal diameter is ≥5.4 cm. RAEF – right atrial emptying fraction and RVID – Right ventricular basal diameter.

Consistent with this, the incident rate ratio (IRR) of AF/AFL in patients with RAEF <17% or RV basal diameter ≥5.4 cm was significantly higher compared to patients with RAEF ≥17% (IRR: 3.43, 95% CI: 1.69 – 6.49, *p*=0.0003) (**Figure 4A**) or RV basal diameter <5.4 cm (IRR: 3.34, 95% CI: 1.87 – 6.15, *p*<0.001) (**Figure 4B**), respectively. The incident rate ratio of AF/AFL in patients with both RAEF <17% and RV basal diameter ≥5.4 cm was even significantly higher (IRR: 6.60, 95% CI: 2.65 – 15.72, *p*<0.001, **Figure 4C**).

**Figure 4.**
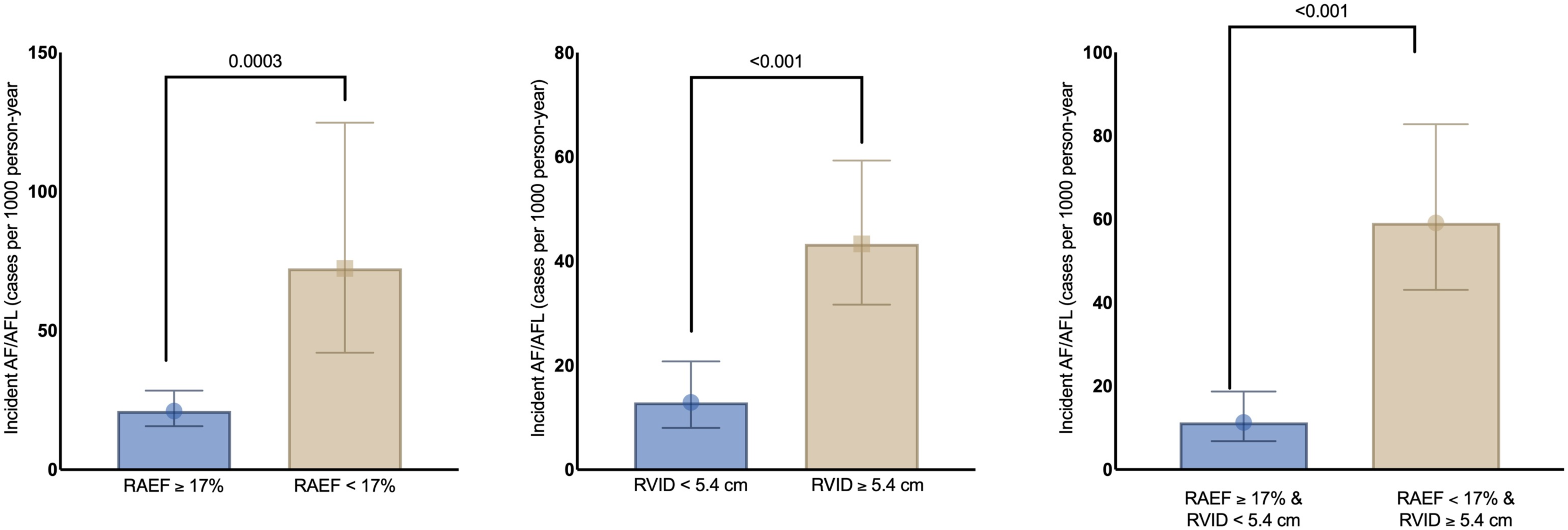
Incidence of Atrial Fibrillation and Atrial Flutter Categorized by Right Atrial Emptying Fraction (RAEF) and Right Ventricular (RV) Basal Diameter (A) PAH patients with RAEF <17% had a significantly higher incidence rate of AF/AFL compared to patients with RAEF ≥17%. (B) PAH patients with RV basal diameter ≥5.4 cm had a significantly higher incident rate of AF/AFL compared to patients with RV basal diameter <5.4 cm. (C) Incident rate ratio of AF/AFL in patients with both RAEF <17% and RV basal diameter ≥ 5.4 cm was significantly higher when compared to those with RAEF ≥17% and RV basal diameter <5.4 cm.

Likewise, on Kaplan-Meier analysis, patients with RAEF <17% or RV basal diameter ≥5.4 cm had significantly lower probability of being free of incident AF/AFL compared to patients with RAEF ≥17% (*p*=0.001, **Figure 5A**) or RV basal diameter <5.4 cm (P=0.006, **Figure 5B**), respectively. Patients with both RV basal diameter ≥ 5.4 cm and RAEF <17% had the lowest probability of being free from incident AF/AFL whereas patients with RV basal diameter <5.4 cm and RAEF ≥17% had the highest probability of being free from incident AF/AFL (P=0.001, **Figure 5C**).

**Figure 5.**
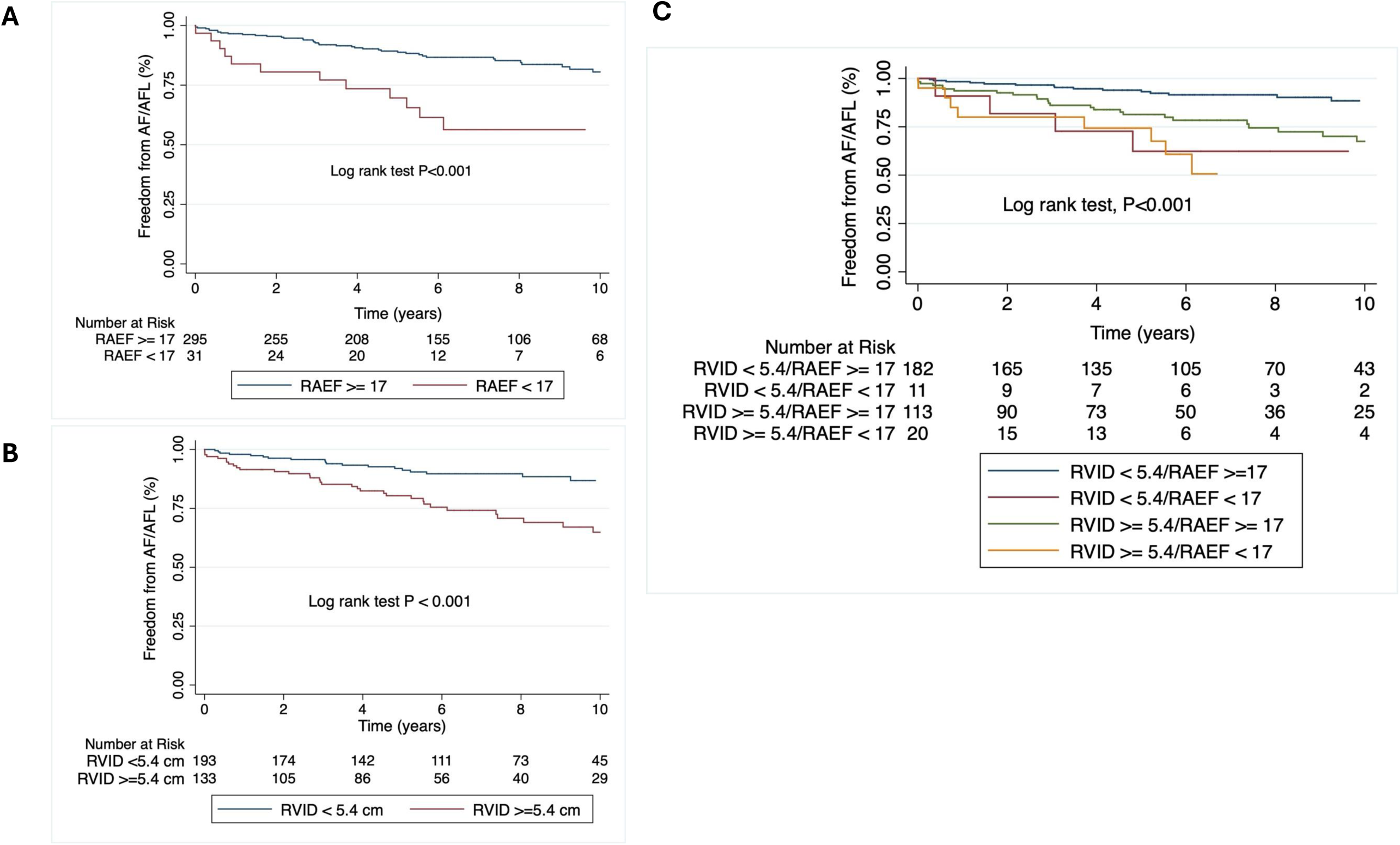
Comparison of Kaplan-Meier Curves for Probability of Incident Atrial Fibrillation and Atrial Flutter Categorized by RAEF, RV Basal Diameter, and Combination of RAEF and RV Basal Diameter. (A) PAH patients with RAEF <17% had significantly lower probability of being free from incident AF/AFL compared to patients with RAEF ≥17%. (B) PAH patients with RV basal diameter ≥5.4 cm had significantly lower probability of being free from incident AF/AFL compared to patients with RV basal diameter <5.4 cm. (C) Patients with both RV basal diameter ≥ 5.4 cm and RAEF <17% had the lowest probability of being free from incident AF/AFL whereas patients with RV basal diameter <5.4 cm and RAEF ≥17% had the highest probability of being free from incident AF/AFL.

## Discussion

In a well characterized and sufficiently sized cohort of PAH patients in the contemporary era, we provide insights into the relationship between right heart remodeling and right atrial function and incident AF/AFL. First, we show the incident rate of AF/AFL in a multicenter cohort of PAH patients with an intermediate risk category REVEAL lite score is 25.3 cases per 1000 person-year. Second, RV dilatation and RA mechanical dysfunction are shown to be independent determinants of incident AF/AFL in PAH patients. Third, we identify cutoff values of RV basal diameter (≥5.4 cm) and RAEF (<17%) that are associated with increased risk of incident AF/AFL in PAH patients. Taken together, these observations could help clinicians develop tailored monitoring and early diagnostic strategies to identify AF/AFL in PAH patients, thereby potentially improving clinical outcomes.

In a single center study of 317 PAH patients, Mercurio et al., report a cumulative incidence of 13.2% AF/AFL over a mean follow up of 67 ± 51 months(7). Wen et al. note a cumulative incidence of 15.8% supraventricular tachycardias that include atrial fibrillation, atrial flutter, and atrial tachycardia after six years(5). In a multicenter study of 297 patients with PAH or CTEPH, Smith et al. observe a cumulative incidence of 26% AF/AFL at some point over the disease course(8). Olsson et al., report a cumulative incidence of 25% AF/AFL over 5 years in 239 patients with either idiopathic PAH or CTEPH(6). These prior studies in PAH patients report only cumulative incidence of AF/AFL over a variable observation period and some of these studies include both patients with PAH and CTEPH(5–8), making it difficult to understand the true rate of incident AF/AFL in PAH.

To address this knowledge gap, we studied only PAH patients and calculated an incident rate of AF/AFL using a time to event analysis, which accounts for potential difference in time to occurrence of AF/AF by incorporating the time at risk. The incident rate of AF/AFL in our prevalent PAH cohort with a mean REVEAL score of 7.4 is 25.3 cases per 1000 person-year. This is higher than the incident rate of 6.82 cases of AF/AFL per 1000 person-year in the general population(10), but much lower when compared to the incidence rate of 231 cases of AF/AFL per 1000 person-year in patients with left heart failure(11). Thus, PAH patients are at increased risk of AF/AFL, but this disease does not carry that same risk as left heart failure. Of the AF/AFL in our cohort, atrial fibrillation is much higher in incidence than atrial flutter (87.5 % vs. 12.5%).

Prior studies evaluating the determinants of AF/AFL in PAH included only demographic, clinical, and hemodynamic characteristics, but did not study RA and RV dimensions and mechanics (6–8). Little is known about the influence of RA and RV remodeling and mechanics on the risk of incident AF/AFL in healthy subjects as well as in PAH patients. In the Atherosclerosis Risk in Communities (ARIC) Study, increased RV afterload (mPAP and PVR) is associated with heightened risk of incident AF independent of left atrial and left ventricular remodeling in healthy volunteers(12). Similarly, higher RA volume index is associated with increased risk of incident AF in healthy subjects in the Multi-Ethnic Study of Atherosclerosis (MESA)(13). Our study aligns with a multicenter study of 280 idiopathic PAH patients by Wen et al., which identified RV basal diameter, mean RA pressure, and left atrial area as independent predictors of AF/AFL and atrial tachycardia(5). Thus, our study systemically defines how right heart remodeling impacts AF/AFL risk in PAH, and parallels findings from other groups showing how cardiac remodeling impacts AF/AFL in broader clinical cohorts.

When comparing our results to Wen et al(5), we find important similarities and differences that require further evaluation. Interestingly, like Wen et al., we find that RV basal diameter is an independent predictor of incident AF/AFL. However, left atrial remodeling as assessed by left atrial volume index is not an independent predictor of incident AF/AFL in our cohort. While mean RA pressure is associated with incident AF/AFL in our univariate analysis, this is no longer significant after adjusting for other covariates in the multivariable analysis. The exact reasons for the discrepancies in our observation when compared to Wen et al. is unclear. However, differences in the patient characteristics and the primary outcome may shed light on our divergent findings. Patients in the study by Wen et al. had more severe PAH compared to our cohort, and their primary outcome included atrial tachycardia in addition to AF/AFL.

Interestingly, our multivariable model demonstrated RV dilatation, defined by RV basal diameter, is the most independent predictor of incident AF/AFL among all the other right heart structural remodeling parameters. While our study does not provide a mechanistic link between RV dilatation and incident AF/AFL, there are several possible reasons. First, RV dilatation leads to tricuspid valve annular dilatation, malcoaptation of the tricuspid valve leaflets, and tricuspid regurgitation(2). Tricuspid regurgitation causes volume overload of the right atrium, which in addition to the pressure overload in the setting of pulmonary vascular disease, causes more adverse RA structural and functional remodeling(14), potentially leading to AF/AFL. Consistent with this, we report a significant relationship between the severity of tricuspid regurgitation and RV basal diameter in our cohort (**Supplemental Figure 2**). One potential reason why RV diameter is a stronger predictor than RA remodeling is that echocardiographic measurements of RA size such as RA area and volume assume that the RA remodels concentrically in a uniform fashion (9). However, in PAH, due to significant pressure and volume overload, the RA enlarges non-uniformly with a higher eccentricity index(14). Thus, RA size and volume assessed by 2DE probably does not accurately describe RA structural remodeling in PAH due to limited ability to capture the nonlinear remodeling. In contrast, RV basal diameter is a simple linear measurement that requires less assumption regarding cardiac anatomy, which heightens its reproducibility and accuracy.

While there is no independent association between RA structural remodeling and the risk of incident AF/AFL, we observe an independent association between RA mechanical dysfunction, as measured by RAEF from 2DE, and increased risk of AF/AFL. While the exact mechanism behind the association between low RAEF and increased risk of incident AF/AFL is unclear, we postulate that low RAEF is more predictive of incident AF/AFL than RA structural remodeling parameters as it is more closely associated with the presence and extent of atrial fibrosis(3). Pressure and volume overload of the RA leads to atrial myopathy characterized by increased atrial fibrosis and capillary rarefaction in PAH(3). RA fibrosis heightens AF inducibility in preclinical models of PAH and in PAH patients by facilitating the formation of reentrant circuits(15,16). In support of our observation, lower left atrial EF or left atrial mechanical dysfunction increases risk of AF/AFL and lowers the likelihood of staying in sinus rhythm after cardioversion, when compared to left atrial structural remodeling in patients with left heart disease(17). Alternatively, it is possible that low RAEF is more predictive of incident AF/AFL as its measurement is less dependent on the angle of the 2DE images compared to other RA and RV echocardiographic measurements. Finally, it is conceivable that low RAEF predicts increased risk of AF/AFL as it is a more robust readout of poor RV function.

Interestingly, both RAEF and RV basal diameter have an additive ability to predict incident AF/AFL. Patients with RAEF <17% and RV basal diameter ≥5.4 cm have a much lower probability of being free from incident AF/AFL when compared to those with either RV basal diameter <5.4 cm or RAEF ≥17% alone. Thus, the presence of both RV basal diameter ≥5.4 cm and RAEF <17% identifies a subset of population who are at a relatively higher risk of incident AF/AFL and therefore should be monitored closely.

Higher REVEAL lite risk score also predicts increased risk of incident AF/AFL (**Supplemental Figure 3**). Of note, when we adjust for REVEAL lite score (as a continuous variable) instead of REVEAL risk status (as a categorical variable – low, intermediate, or high risk) in our multivariable model, RAEF is no longer significantly associated with incident AF/AFL. Only RV basal diameter and REVEAL lite score are independently associated with incident AF/AFL. This is likely because REVEAL lite score is a multiparametric global measure of right heart function.

## Limitations

Although both the Minnesota Pulmonary Hypertension repository and the Stanford Pulmonary Hypertension registry are prospective observational cohorts, we collected data on incident AF/AFL retrospectively. Thus, we included only clinically identified incident AF/AFL. We did not do continuous remote monitoring or routine, periodic, surveillance electrocardiograms. Hence, it is possible that some patients who might have had asymptomatic AF/AFL were missed. This could have led to underestimation of the burden of incident AF/AFL in PAH. Second, we used 2DE to assess RA structural remodeling and mechanical dysfunction. However, cardiac magnetic resonance imaging (MRI) provides relatively more accurate measurement of RA size and deeper characterization of RA mechanical dysfunction than 2D echocardiography. Nevertheless, 2D is universally available and routinely done as opposed to cardiac MRI, which is used sporadically in the management of PAH. Thus, our data may be more relevant for clinical practice. We studied only incident AF and AFL but did not include atrial tachycardias. Finally, we included both prevalent and incident patients. The incidence of AF/AFL in PAH may vary in incident compared to prevalent PAH cohorts as well as be affected by severity of PAH.

## Conclusions

In conclusion, we report incident rates of AF/AFL in a multicenter cohort of PAH patients with mean REVEAL lite score in the intermediate risk category. We further demonstrate the importance of RV basal diameter and RAEF to identify patients with PAH who are at increased risk of incident AF/AFL.

## Data Availability

All data produced in the present study are available upon reasonable request to the authors

---

**Supplemental Figure 1.**
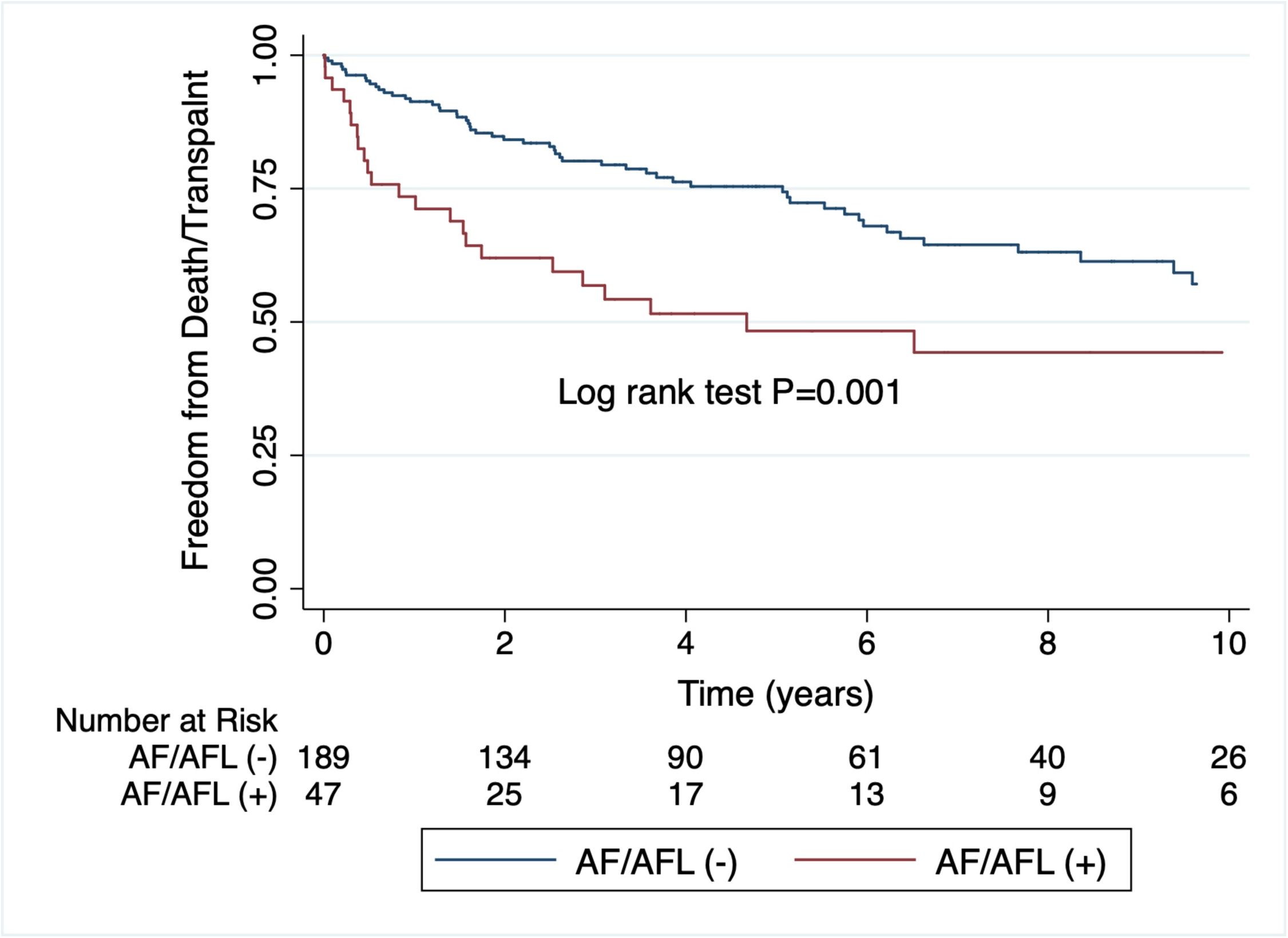
Comparison of Survival Free of Death and Transplantation Categorized by Incident Atrial Fibrillation and Flutter. Patients who developed incident AF/AFL experienced significantly higher rates of death or transplantation compared to those who did not. For patients with incident AF/AFL, the date of AF/AFL onset was used as the entry point for the survival analysis. In contrast, for those without incident AF/AFL, the survival analysis began at the date of diagnosis + median time to onset of incident AF/AFL (3.1 years) to make the two groups more comparable.

**Supplemental Figure 2.**
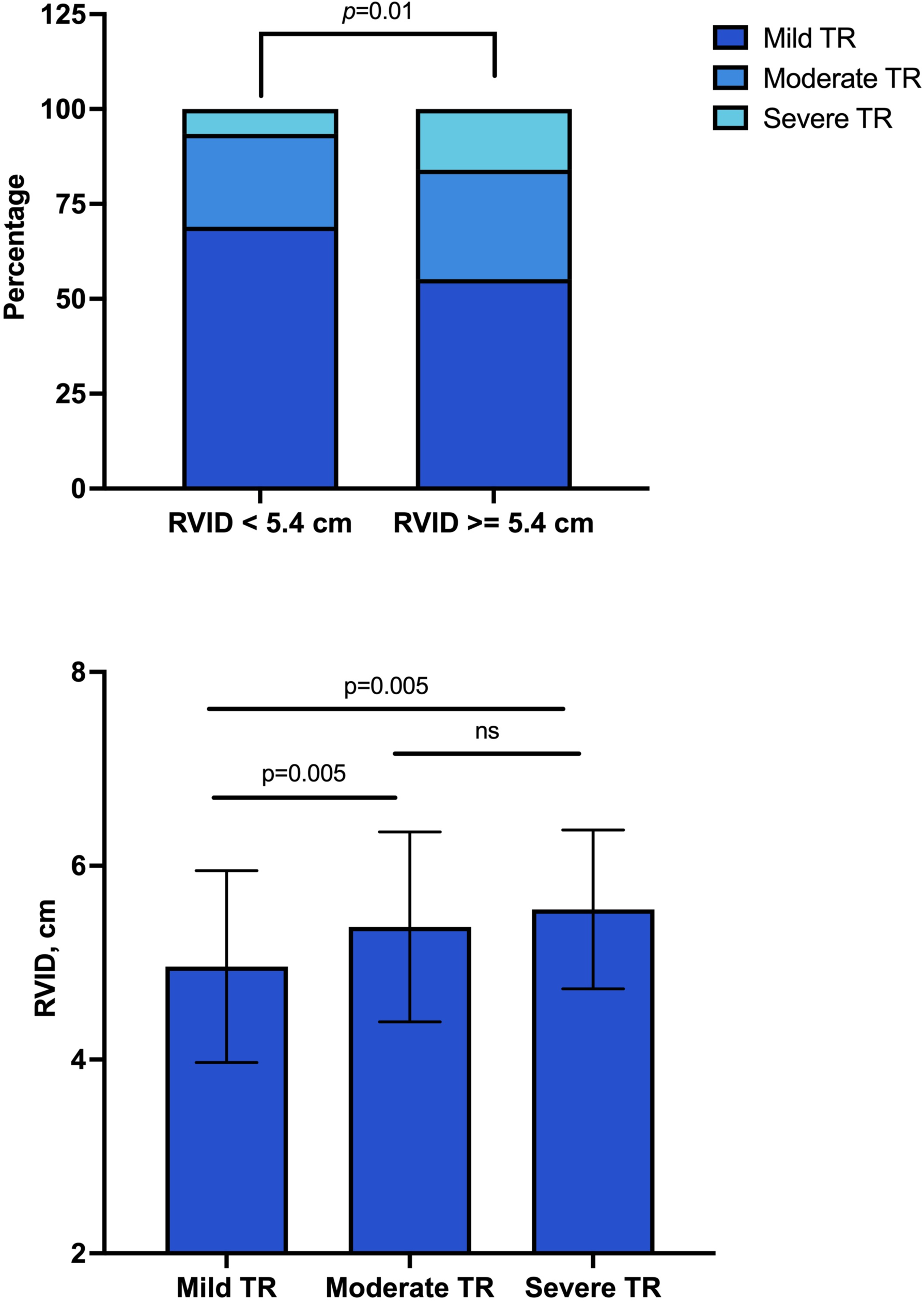
Relationship Between Right Ventricular Basal Diameter and Tricuspid Regurgitant Velocity. PAH patients with RV basal diameter ≥ 5.4 cm had significantly higher number of patients with moderate or severe tricuspid regurgitation when compared to those with RV basal diameter < 5.4 cm (Top panel). Likewise, RV basal diameter is significantly higher in PAH patients with moderate or severe tricuspid regurgitation as compared to those with mild tricuspid regurgitation. RV – right ventricle and TR – tricuspid regurgitation (Bottom panel).

**Supplemental Figure 3.**
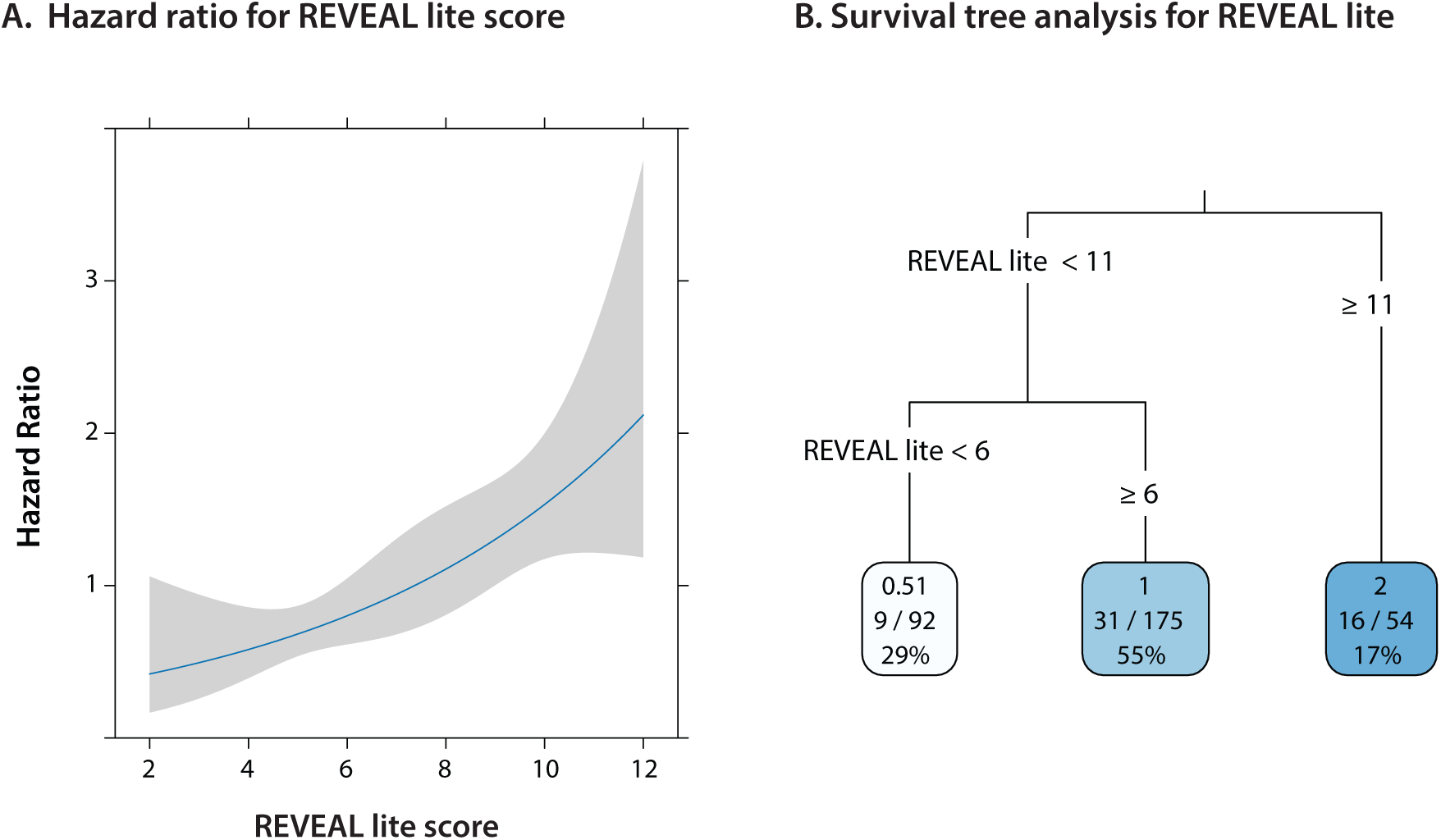
Non-linear Restricted Cubic Spline Analyses (A) and Survival Decision Tree Analysis (B) for Identifying Threshold of REVEAL Lite score Associated with Incident Atrial Fibrillation and Flutter in PAH (A) The risk of incident AF/AFL increases when the REVEAL lite score is ≥7.4. (B) The risk of incident AF/AFL increases when the REVEAL lite score is ≥6, but the risk increases significantly when the REVEAL lite score is ≥11. REVEAL -- Registry to evaluate early and long-term PAH disease management.

